# Food insecurity disparities and mental health impacts among cancer survivors during the COVID-19 pandemic

**DOI:** 10.1101/2022.02.06.22270283

**Authors:** Marlene Camacho-Rivera, Jessica Yasmine Islam, Diane R. Rodriguez, Denise C. Vidot, Zinzi Bailey

## Abstract

Food insecurity can negatively impact adherence and receipt of high-quality cancer care. The purpose of the study was to (1) compare the prevalence of COVID-19 associated food insecurity by cancer history and (2) examine determinants associated with COVID-19 related food insecurity among cancer survivors.

We used nationally-representative data from the COVID-19 Household Impact Survey (n = 10,760), collected at three time points: April 20-26, May 4-10, and May 30th -June 8^th^ of 2020. Our primary exposure was cancer survivor status, based on participant’s self-report of a cancer diagnosis (n=854, 7.1%). Primary outcomes of food insecurity were categorized on how often participants reported the following: “We worried our food would run out before we got money to buy more” or “The food that we bought just didn’t last, and we didn’t have money to get more”; Respondents were categorized as food insecure if they chose often true or sometimes true. Multivariable Poisson regression was used to identify demographic determinants of food insecurity among cancer survivors.

Thirty-two percent of cancer survivors were food insecure. Cancer survivors aged 30-44 years and those aged ≥60 were more likely to report being food insecure compared to respondents without a history of cancer in the same age categories (30-44 years, 59.9% versus 41.2% *p* = 0.01, ≥60 years 27.2% versus 20.2%, *p* = 0.01). Cancer survivors without a high school diploma were more likely to report food insecurity compared to adults with no history of cancer (87.0% versus 64.1%, *p* = 0.001). In multivariable models, uninsured cancer survivors (adjusted Prevalence Ratio (aPR) aPR: 2.39, 95% CI: 1.46-3.92) and those on Medicaid (aPR: 2.10, 95% CI: 1.40-3.17) were also more likely to report being food insecure.

Food insecurity during the COVID-19 pandemic is vast but disparities persist. Among cancer survivors, differences in food insecurity were observed by age and SES.

## Introduction

The COVID-19 pandemic and resulting lockdowns has caused widespread economic, social, and public health effects. One such consequence of the pandemic has been an increase in households facing food insecurity in the US (1). The United Nations defines food security as when people “have physical and economic access to sufficient safe and nutritious food that meets their dietary need requirement and food preferences” (2). They further define four dimensions that must be met to be food secure, including physical availability of food, economic and physical access to food, food utilization, and the stability of these factors over time (2). According to the USDA annual report in 2018, 11.1% of households in the US were food insecure at any point during the year (3). Food insecurity is more common in urban areas, immigrant and ethnic minority populations, and households that earn below 185% of the federal poverty line (3,4). Food insecurity parallels economic conditions such as unemployment rates and poverty (1,5). During periods of economic downturn, there is a corresponding increase in food insecurity (5). For example, during the recession in the United States that began in 2008, there was an increase in food insecurity, increased usage of food pantries, and an increase in unemployment (6).

Food insecurity is a threat to public health; patients who screen positive for food insecurity have higher risk of adverse health outcomes, including diabetes, hypertension, mental illness, and increased mortality (1,7). Food insecurity is also associated with significantly more emergency department visits and hospitalizations with subsequently higher healthcare expenditures (8). In a Canadian study of households facing food insecurity, these populations had a significantly higher rate of anxiety and mental health symptoms during the COVID-19 pandemic than households that were food secure (9). In addition to higher rates of psychological symptoms, food insecurity can weaken the body’s immune system, making an individual more susceptible to infection (10).

The COVID-19 pandemic has disrupted all four factors that contribute to food security. Millions of Americans lost their jobs in the months following lockdowns and social distancing measures, resulting in new or worsening economic barriers to food security. Public transportation was disrupted with decreased access in efforts to maintain social distancing, presenting a physical barrier to obtaining food (1). Additionally, millions of Americans stayed home in efforts to decrease the spread of COVID-19. In a study of the early effects of COVID-19, a web based survey was administered to adults who earned less than 250% of the federal poverty line between 19 March and 24 March 2020, when lockdowns and social distancing efforts were just beginning to take place (11). In this study, 44% were food insecure, and an additional 20% had marginal food security (11). A population-level survey in Vermont demonstrated a 32% increase in household food insecurity in the first two weeks of April 2020, with 35% of food insecure households being newly food insecure (1). The economic factors, social distancing measures, and social determinants in the wake of the COVID-19 pandemic make vulnerable populations especially susceptible to food insecurity (12).

Cancer patients and those with a history of cancer are vulnerable to food insecurity, especially if they are an underserved population (13-15). Food insecurity may be prevalent in cancer patients for several reasons including high cost of medical care, changes in employment or income after cancer diagnosis, or other causes of financial strain (13,16). Food insecurity in cancer patients is associated with worse quality of life and psychological wellbeing (15). In a study of over 1300 underserved patients receiving cancer care in New York City, 41% had low food security, and 17% had very low food security (13). In another study of urban low income cancer patients, 56% were found to be food insecure (4). Younger age, primarily Spanish speaking, and not having a primary care provider were factors associated with a greater degree of food insecurity in cancer patients (4).

The COVID-19 pandemic has had wide ranging effects on the US economy and food insecurity in vulnerable populations. Cancer patients are susceptible to financial strain and food insecurity due to cancer treatment cost, loss of employment, and competing demands. Therefore, those with a current or past history of cancer diagnosis may be especially vulnerable to the economic effects of the pandemic. So far no studies have examined the economic effects of the COVID-19 pandemic specifically on those with a current or past history of cancer, who are a uniquely vulnerable population. The aim of this study was to examine the effect of the COVID-19 pandemic on food insecurity in patients with a history of cancer.

## Methods

### COVID-19 Impact Survey

We used publicly available data from the COVID-19 Impact survey, a weekly survey administered to a nationally representative sample of 10,760 adults in the United States by the NORC at University of Chicago for the Data Foundation (17). The survey was administered starting in April 2020, with the aim to assess physical, mental, and economic health in the US (17). For this analysis, we leveraged the national data collected during Week 1 (April 20-26, 2020), Week 2 (May 4-10, 2020), and Week 3 (May 30^th^-June 8^th^, 2020), which were merged for this analysis.

### AmeriSpeak Sample

Funded and operated by NORC at the University of Chicago, AmeriSpeak® is a probability-based panel designed to be representative of the US household population. During the initial recruitment phase of the AmeriSpeak panel, randomly selected US households were sampled using area probability and address-based sampling, with a known, nonzero probability of selection from the NORC National Sample Frame. These sampled households were then contacted by US mail, telephone, and field interviewers (face to face). The panel provides sample coverage of approximately 97% of the US household population. Those excluded from the sample include people with P.O. Box only addresses, some addresses not listed in the US Postal Service Delivery Sequence File, and some newly constructed dwellings. While most AmeriSpeak households participate in surveys by web, non-internet households were able to participate in AmeriSpeak surveys by telephone. Households without conventional internet access but having web access via smartphones could participate in AmeriSpeak surveys by web. AmeriSpeak panelists participate in NORC studies or studies conducted by NORC on behalf of governmental agencies, academic researchers, and media and commercial organizations. Interviews were conducted in English and Spanish. Interviews were conducted with adults aged 18 years and above and over representing the 50 states and the District of Columbia. Panel members were randomly drawn from AmeriSpeak. In households with more than one adult panel member, only one was selected at random for the sample. Invited panel members were given the option to complete the survey online or by telephone with an NORC telephone interviewer. The number of participants invited, and percentage of interviews completed by week are as follows: 11,133 invited with 19.7% interviews completed during Week 1; 8,570 invited with 26.1% interviews completed (Week 2); and 10, 373 invited with 19.7% interviews completed (Week 3). The analytic sample includes 10,760 adults nationwide. The final analytic sample were weighted to reflect the US population of adults aged 18 years and over. The demographic weighting variables were obtained from the 2020 Current Population Survey.

### Cancer Survivors

We defined cancer survivors as those participants with a self-reported cancer diagnosis. Participants were asked the following question: “Has a doctor or other health care provider ever told you that you have any of the following: Diabetes; High blood pressure or hypertension; Heart disease, heart attack or stroke; Asthma; Chronic lung disease or COPD; Bronchitis or emphysema; Allergies; a Mental health condition; Cystic fibrosis; Liver disease or end-stage liver disease; Cancer; a Compromised immune system; or Overweight or obesity.” We defined those who selected “Cancer” as a cancer survivor, similar to our previously published work.

### Primary Outcomes – Food Insecurity and Mental Health Symptoms

Our primary measures for this analysis were food insecurity and mental health symptoms. We defined food insecurity using the following questions: Please indicate whether the following statements were often true, sometimes true, or never true over the past 30 days: “We worried our food would run out before we got money to buy more.” “The food that we bought just didn’t last, and we didn’t have money to get more.” Participants who reported often true or sometimes true to either of the two previous statements were categorized as reporting food insecurity. Participants were also asked to self-report whether, in the past 7 days, they had either received, applied for, tried to apply for, or did not receive nor apply for any of the following income assistance: Food pantry assistance or Supplemental Nutrition Assistance Program(SNAP). Participants who had reported receiving or trying to apply for food pantry assistance or SNAP were considered food insecure.

Next, to evaluate mental health symptoms, participants were asked to report symptoms of anxiety, depression, loneliness, hopelessness, and physical reaction to experiences during the COVID-19 pandemic in the seven days before survey administration. Participants were able to choose from the following list of options for each mental health symptom: Not at all or less than 1 day, 1-2 days, 3-4 days, 5-7 days. For multivariable analyses, we categorized the frequency of mental health symptoms as follows: Not all or less than one day, 1-2 days per week, and 3-7 days per week due to limited sample size.

### Covariates

The following covariates were included in the multivariable analyses: age (18-29, 30-44, 25-59, 60+), gender (male/female), marital status (married/living with a partner, widowed/divorced/separated, never married), race/ethnicity categories [non-Hispanic (NH) White, NH-Black, Hispanic, NH-Asian, NH-Other], education categories (no high school diploma, HS graduate or equivalent, some college, baccalaureate degree or above), employment status (employed/unemployed), household income (<$50,000, $50,000-<$100,000, ≥$100,000), population density (rural, suburban, urban), census region (Northeast, Midwest, South, West), and insurance type (purchased plan/employer-sponsored/TRICARE/Medicaid/Medicare/Dually-eligible/VA/uninsured).

### Data Analysis

Descriptive statistics are summarized, by cancer survivorship status, in percentages among all respondents and include a margin of error of +/-3·0 percentage points at the 95% confidence intervals. We used chi-square tests to compare the prevalence of food insecurity among cancer survivors compared to adults without a history of cancer by key demographic covariates. To identify demographic groups that may be more likely to report food insecurity, we estimate determinants of food insecurity among cancer survivors. We computed prevalence ratios with Poisson regression using robust estimation of standard errors.(12–14) Potential variables for inclusion in the model were assessed using available sociodemographic variables and bivariate Poisson regression analysis. Due to the exploratory nature of this analysis using a predictive framework, an arbitrary p-value of <0.10 was used as criteria to include the variable in the multivariable Poisson regression model. For multivariable Poisson regression models, adjusted prevalence ratios (aPR), and 95% confidence intervals (CIs) for each independent variable were calculated.

Next, we used multinomial logistic regression to evaluate associations between food insecurity and mental health symptoms reported in the last 7 days among cancer survivors. We adjusted for age, sex, race/ethnicity, annual household income, education, insurance status, employment status, and area of residence (urban/rural). To address concerns regarding existing mental health symptoms before the COVID-19 pandemic, we conducted a sensitivity analysis to evaluate mental health symptoms among those without a history of a mental health condition based on self-report. We were able to assess the history of a mental health condition through the following question: “Have you ever been diagnosed by a doctor or health care provider ever said you have a mental health condition?” Although “mental health condition,” may include several conditions, we were able to focus on those without clinical depression and anxiety using this approach. Based on the exploratory nature of this analysis, we did not include an adjustment for multiple comparisons(15,16). All statistical analyses were conducted using Stata IC 15 (StataCorp LLC, College Station, TX). Sampling weights were applied to provide results that were nationally representative of the U.S. adult population.

## Results

There were 10,720 adult participants in the COVID-19 impact survey (table 1). To the item “over the past 30 days, we worried our food would run out before we got money to buy more” 6.2% (5.5,6.9) answered “Often true,” 20% (18.9,21.2) answered “sometimes true,” with no significant difference between cancer survivors and non cancer survivors (table 2). Overall, the prevalence of food insecurity decreased with increasing age, with 47.3% of those age 18-29 reporting food insecurity, and 21% of those over age 60 reporting food insecurity. The highest rates of food insecurity were reported in individuals who self-identified as non-Hispanic Black (50.9%) or Hispanic (49.6%), and lowest among non-Hispanic White participants (25.6%). Participants not currently employed were more likely to be food insecure (39.8%) than employed participants (27.5%). More education was associated with decreased prevalence of food insecurity. Those who reported not having a high school diploma were most likely to be food insecure (65.3%), while the least likely had a baccalaureate degree or above (16%). Other demographic factors associated with the highest prevalence of food insecurity includes household income less than $30,000 (63.4%), living in a rural area (36.5%), and having Medicaid (66.8%) or Indian Health Service insurance (82.5%).

**Table 1.**
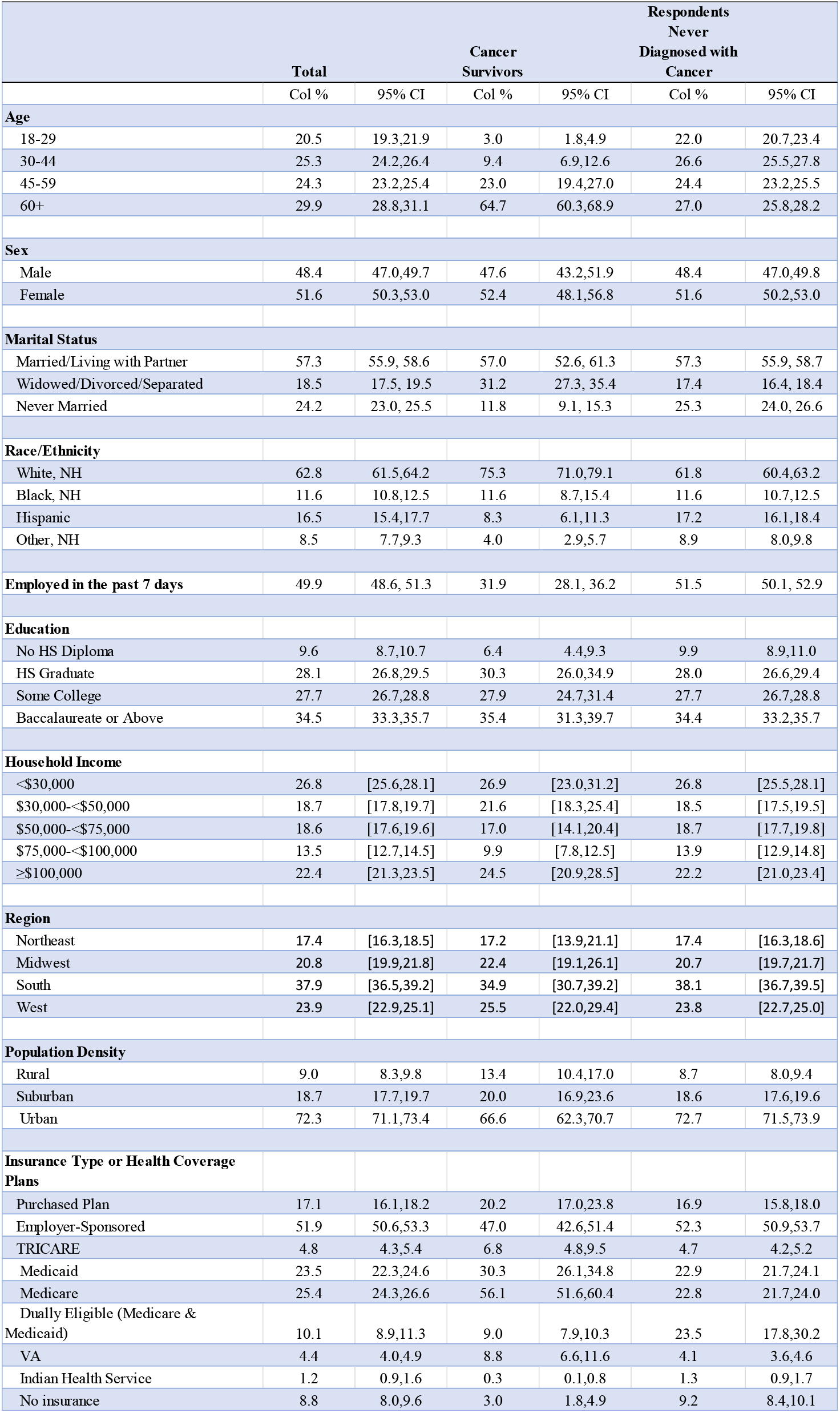
Descriptive characteristics of COVID-19 Impact Survey respondents by cancer survivorship status (n=10,760)

**Table 2.** Prevalence of food insecurity by cancer survivorship status (n=10,760)

| **Food Insecurity Measures** | **Total** |  | **Cancer survivors** |  | **No cancer** |  |
| --- | --- | --- | --- | --- | --- | --- |
| **Over the past 30 days, we worried our food would run out before we got money to buy more** |  |  |  |  |  |  |
| Often true | 6.2 | 5.5,6.9 | 5.2 | 3.4,8.0 | 6.3 | 5.6,7.0 |
| Sometimes true | 20.0 | 18.9,21.2 | 17.5 | 14.0,21.8 | 20.2 | 19.0,21.5 |
| Never true | 73.8 | 72.6,75.0 | 77.2 | 72.8,81.2 | 73.5 | 72.2,74.8 |
| **Over the past 30 days, the food that we bought just didn't last, and we didn't have money to get more** |  |  |  |  |  |  |
| Often true | 4.5 | 4.0,5.1 | 2.9 | 1.6,5.2 | 4.7 | 4.1,5.3 |
| Sometimes true | 16.7 | 15.6,17.8 | 15.9 | 12.4,20.1 | 16.8 | 15.7,17.9 |
| Never true | 78.8 | 77.6,79.9 | 81.2 | 76.8,84.9 | 78.6 | 77.3,79.8 |
| **In the past 7 days, have you received, applied for any of the following forms of income assistance, or not?: SNAP (Supplemental Nutrition Assistance Program)** |  |  |  |  |  |  |
| Received | 10.7 | 9.9,11.5 | 12.3 | 9.4,15.9 | 10.5 | 9.8,11.4 |
| Applied for | 2.2 | 1.8,2.8 | 0.8 | 0.3,1.7 | 2.4 | 1.9,3.0 |
| Tried to apply for | 1.9 | 1.5,2.2 | 1.0 | 0.5,2.2 | 1.9 | 1.6,2.3 |
| Did not receive nor apply for any | 85.2 | 84.3,86.2 | 85.9 | 82.2,89.0 | 85.2 | 84.2,86.1 |
| **In the past 7 days, have you received, applied for any of the following forms of income assistance, or not?: A food pantry** |  |  |  |  |  |  |
| Received | 6.7 | 6.1,7.4 | 7.9 | 5.8,10.7 | 6.6 | 6.0,7.3 |
| Applied for | 0.9 | 0.6,1.2 | 0.9 | 0.3,2.1 | 0.9 | 0.6,1.3 |
| Tried to apply for | 1.0 | 0.8,1.3 | 0.0 | 0.0,0.1 | 1.1 | 0.8,1.5 |
| Did not receive nor apply for any | 91.4 | 90.6,92.1 | 91.2 | 88.3,93.4 | 91.4 | 90.6,92.2 |

Among Hispanic participants, there was a significant increase in prevalence of food insecurity in non-cancer survivors (50.6%) when compared to cancer survivors (24.2%, p<0.01). In participants with no high school diploma, cancer survivors were more likely to be food insecure (87%) than non-cancer survivors (64.1%, p<0.01). In those who reported some college, however, respondents never diagnosed with cancer reported a significantly higher rate of food insecurity (35.6%) compared to cancer survivors (24.9%, p<0.001). In participants with no health insurance, 80.7% of cancer survivors were food insecure, compared to 56.5% of those who were never diagnosed with cancer (56.5%, p=0.01).

Within the cancer survivor group (n=854), there were several factors that were independently associated with an increased risk of food insecurity (table 3). Cancer survivors with health insurance through Medicaidor no insurance had higher prevalence ratios of financial insecurity (PR 2.1and 2.39, respectively). Those with no high school diploma or a high school level of education had higher risk of food insecurity than those with baccalaureate or above (PR 2.63 and 1.94, respectively). Household income <$30,000 was associated with higher risk of food insecurity than income over $100,000 (PR2.16; 95% CI 1.15-4.07). Living in a rural area was also associated with a higher risk of financial insecurity than living in an urban area (PR 1.51; 95% CI 1.07-2.12).

**Table 3.** Determinants of food insecurity among cancer survivors (n=853)

|  | Unadjusted<br>PR | 95% CI | Adjusted PR | 95% CI |
| --- | --- | --- | --- | --- |
| **Age** |  |  |  |  |
| 18-29 | 1.90 | 1.12-3.24 | 1.31 | 0.76-2.27 |
| 30-44 | 2.20 | 1.61-3.01 | 1.42 | 1.04-1.95 |
| 45-49 | 1.16 | 0.82-1.64 | 1.54 | 1.16-2.03 |
| 60+ | Ref. |  | Ref. |  |
| **Sex** |  |  |  |  |
| Male | Ref. |  | Ref. |  |
| Female | 1.47 | 1.11-1.94 | 1.13 | 0.88-1.46 |
| **Marital Status** |  |  |  |  |
| Married/Living with Partner | Ref. |  | Ref. |  |
| Widowed/Divorced/Separated | 1.20 | 0.89-1.62 | 0.90 | 0.70-1.15 |
| Never Married | 1.78 | 1.25-2.54 | 1.00 | 0.71-1.42 |
| **Race/Ethnicity** |  |  |  |  |
| White, NH | Ref. |  | Ref. |  |
| Black, NH | 1.82 | 1.29-2.56 | 1.00 | 0.74-1.34 |
| Hispanic | 0.84 | 0.45-1.52 | 0.88 | 0.52-1.48 |
| Asian, NH | 1.62 | 0.71-3.70 | 1.90 | 0.41-8.92 |
| Other, NH | 1.12 | 0.67-1.87 | 1.08 | 0.79-1.49 |
| **Insurance Type\*** |  |  |  |  |
| Purchased Plan | 0.69 | 0.47-1.01 | 1.16 | 0.77-1.74 |
| Employer-Sponsored | 0.50 | 0.37-0.67 | 1.16 | 0.73-1.83 |
| TRICARE | 1.09 | 0.64-1.85 | - |  |
| Medicaid | 3.24 | 2.48-4.23 | 2.10 | 1.40-3.17 |
| Medicare | 0.92 | 0.69-1.21 | - |  |
| Dually Eligible (Medicare & Medicaid)† | 2.25 | 1.74-2.91 | - |  |
| VA | 0.90 | 0.55-1.47 | - |  |
| Indian Health Service | 2.60 | 1.92-3.54 | 2.26 | 0.87-5.89 |
| No insurance | 2.65 | 2.10-3.34 | 2.39 | 1.46-3.92 |
| **Employment Status** |  |  |  |  |
| Not Employed | Ref. |  | Ref. |  |
| Employed/Self-Employed | 0.60 | 0.43-0.83 | 0.83 | 0.57-1.22 |
| **Education** |  |  |  |  |
| No HS Diploma | 6.30 | 4.17-9.40 | 2.63 | 1.53-4.50 |
| HS Graduate | 3.46 | 2.23-5.36 | 1.94 | 1.08-3.49 |
| Some College | 1.79 | 1.15-2.81 | 1.36 | 0.82-2.27 |
| Baccalaureate or Above | Ref. |  | Ref. |  |
| **Household Income** |  |  |  |  |
| <\$30,000 | 4.93 | 2.78-8.74 | 2.16 | 1.15-4.07 |
| \$30,000-<\$50,000 | 2.32 | 1.24-4.34 | 1.48 | 0.74-2.96 |
| \$50,000-<\$75,000 | 1.31 | 0.64-2.59 | 0.99 | 0.52-1.91 |
| \$75,000-<\$100,000 | 1.10 | 0.48-2.54 | 0.97 | 0.47-2.03 |
| ≥\$100,000 | Ref. | | Ref. | |
| **Region** |  |  |  |  |
| Northeast | Ref. |  | Ref. |  |
| Midwest | 0.80 | 0.53-1.19 | 0.78 | 0.52-1.19 |
| South | 0.88 | 0.60-1.28 | 0.67 | 0.46-0.99 |
| West | 0.55 | 0.35-0.86 | 0.73 | 0.46-1.16 |
| **Population Density** |  |  |  |  |
| Rural | 1.77 | 1.28-2.44 | 1.51 | 1.07-2.12 |
| Suburban | 1.06 | 0.75-1.50 | 1.05 | 0.76-1.46 |
| Urban | Ref. |  | Ref. |  |

Being a cancer survivor with food insecurity increased risk of mental health symptoms compared to those who were food secure (Figure 1). Cancer survivors facing food insecurity had an odds ratio of 9.41 (4.56, 19.4) of feeling nervous or anxious 3-7 days per week, and 1.91 (1.02,3.55) for 1-2 days per week. Cancer survivors who were food insecure had higher odds ratios of feeling depressed (6.21, 95% CI 3.19-12.11), lonely (4.65, 95% CI 2.26-9.55), or hopeless (2.65, 95% CI 1.38-5.08) for 3-7 days per week compared to cancer survivors who were not food insecure. Cancer survivors who were food insecure also had higher odds ratio of feeling lonely 1-2 days per week (2.21, 95% CI 1.23-3.95) when compared to food secure cancer survivors.

**Figure 1.**
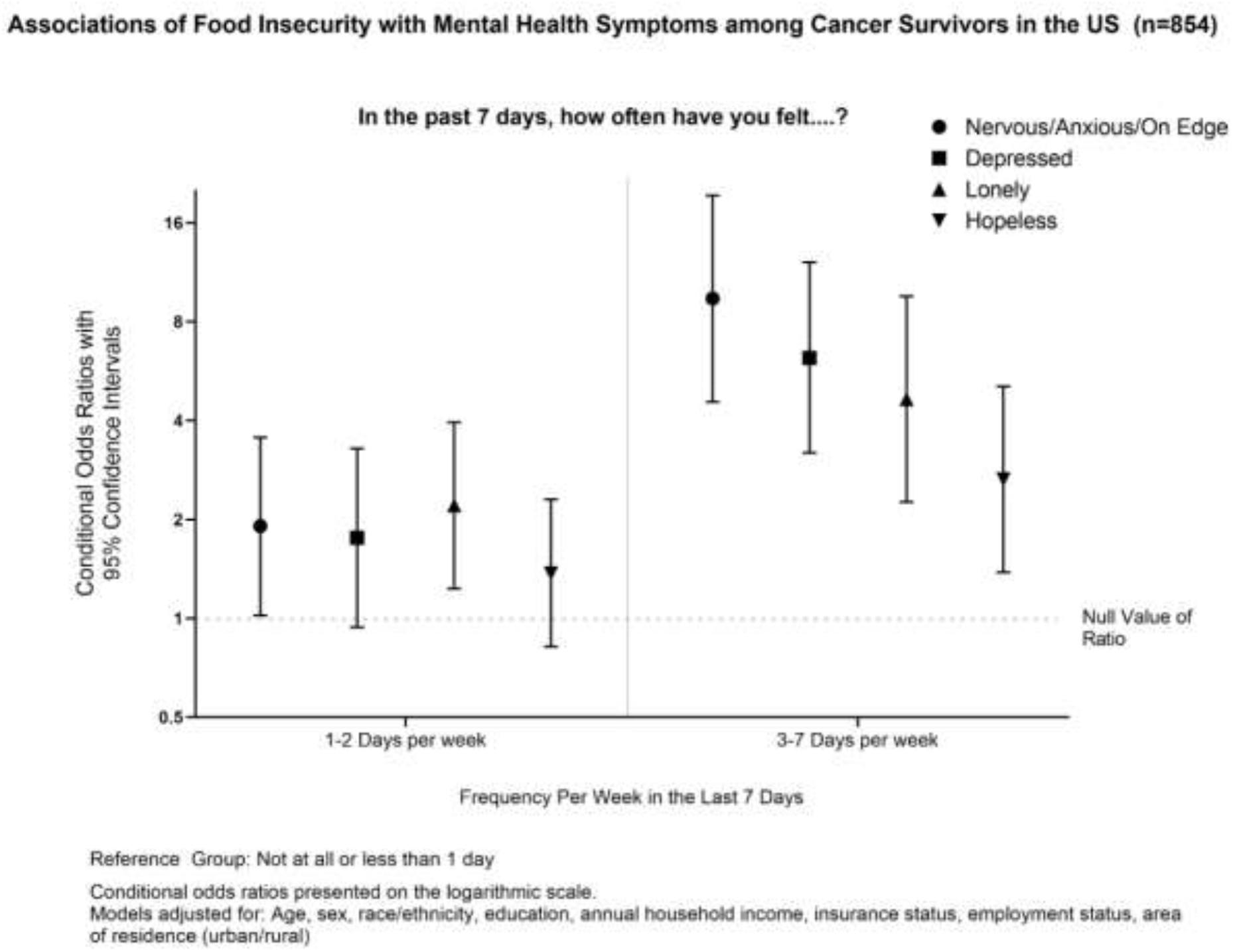

## Discussion

In this study, there was a high prevalence of food insecurity reported in the immediate aftermath of the COVID-19 pandemic, from April-June 2020, when social distancing measures were taking place and millions of Americans became unemployed. 26% of respondents overall reported that in the last month they worried food would run out at least some of the time and 21% said the food they bought did not last and they did not have money to buy more. This compares to the USDA annual report in 2019 that showed 11% of households in the US faced food insecurity at any point during the year (3). The factors associated with an increased risk of food insecurity within the group of cancer survivors mirrored those in the general population and includes mid age categories (ages 30-44 years and 45-49 years), lower levels of education, household income of less than $30,000 per year, residing in a rural area, and having Medicaid (compared to non-Medicaid) or no insurance coverage (compared to any insurance). Each of these factors independently increased risk of reporting food insecurity. This aligns with previous studies showing that these factors increase an individual’s risk of facing food insecurity (7). Minority populations and those with low income are vulnerable populations who have been disproportionately affected by the COVID-19 pandemic. They have higher risk of infections as well as higher risk of facing job loss and financial insecurity.

The goal of this study was to assess whether cancer survivors had a higher risk of facing food insecurity in the immediate aftermath of the pandemic than those never diagnosed with cancer. Within the age groups of 30-44 and over 60, cancer survivors had significantly higher rates of food insecurity than those never diagnosed with cancer. Cancer survivors with no high school diploma as well as those with no health insurance also faced significantly higher rates of food insecurity than those never diagnosed with cancer.

On the other hand, within the Hispanic population and within the group that has some college education, those never diagnosed with cancer faced significantly higher rates of food insecurity than cancer survivors. Among individuals that have some college education, there could have been differences in the distribution of those enrolled in college by cancer history. Although are unable to ascertain whether those with some college education were currently enrolled in colleges or universities, there is emerging evidence of the burden of food insecurity among college students during the COVID-19 pandemic (18-24). Hispanic survey respondents were generally younger than other racial/ethnic groups, with only 17% over the age of 60 years, compared to NH-White adults (35% over the age of 60 years) and NH-Black adults (30% over the age of 60 years). Due to their younger age distribution, the proportion of Hispanic adults with cancer was also lower than other racial/ethnic categories (3% compared to 9% of NH-White adults and 8% of NH-Black adults). Although we were unable to account for household composition within the study, Hispanic adults without a history of cancer may have had more children within the household. Other nationally representative surveys have found the highest burden of food insecurity among households with children (25, 26).

This study also examined what factors within the cancer survivor population were the greatest determinants of financial insecurity. Having Medicaid or no insurance had a prevalence ratio over 2 of food insecurity compared to those with health insurance. Education with less than a high school diploma was also associated with a food insecurity prevalence ratio of 2.63 compared to baccalaureate or above. Household income less than $30,000 also had an adjusted prevalence ratio over 2 compared to over $100,000. Cancer survivors are vulnerable to facing financial difficulties and food insecurity, and these factors within this group further increase their risk of facing these hardships.

Food insecurity is associated with a worse quality of life. Cancer survivors facing food insecurity were 9 times more likely to feel nervous or anxious 3-7 days per week than those who were food secure. They were also much more likely to feel depressed, lonely, or hopeless than those who were food secure, and especially so on more days of the week, as shown in figure 1.

There are several limitations in this study. Cancer survivors were defined as anyone with a current or past history of cancer and did not differentiate those who currently are undergoing cancer treatment and those who may have a more remote history of cancer. Additionally, we did not differentiate between different subtypes of cancer, which may confer different risks of financial insecurity. All of the data including demographics, cancer survivorship status, measures of food insecurity, and mental health symptoms were self-reported.

Cancer survivors face a high level of food insecurity compared to the general population, due to job loss and the financial impact of cancer care. Food insecurity is also associated with worse quality of life and increased risk of adverse health outcomes; therefore, it is an important factor in public health. This study demonstrated that cancer survivors with food insecurity had drastically higher prevalence of mental health symptoms than those who did not face food insecurity.

The COVID-19 pandemic disproportionately affected populations that were already vulnerable to food insecurity, compounding a problem that already existed in the United States. There has been a drastic increase in unemployment in the US as well as the population of households facing economic hardship. In patients newly diagnosed with cancer, screening for financial and food security could be an important part of their cancer care. In other patients, healthcare providers can screen for food insecurity and provide resources when necessary. In this way, healthcare providers can play an important role in addressing disparities that make populations more vulnerable to the economic effects of the pandemic.

## Data Availability

All data produced are available online at https://www.norc.org/Research/Projects/Pages/covid-impact-survey.aspx.

